# Enhancing HIV testing yield in southern Mozambique: the effect of a Ministry of Health training module in targeted provider-initiated testing and counselling

**DOI:** 10.1101/2023.09.25.23296036

**Authors:** Anna Saura-Lázaro, Sheila Fernández-Luis, Tacilta Nhampossa, Laura Fuente-Soro, Elisa López-Varela, Edson Bernardo, Orvalho Augusto, Teresa Sánchez, Paula Vaz, Stanley C. Wei, Peter Kerndt, Nely Honwana, Peter Young, Guita Amane, Fernando Boene, Denise Naniche

**Affiliations:** ISGlobal, Hospital Clínic, Universitat de Barcelona, Barcelona, Spain; Centro de Investigação em Saúde de Manhiça (CISM), Maputo, Mozambique; Instituto Nacional de Saúde (INS), Maputo, Mozambique; Manhiça District Health Services, Maputo Province, Mozambique; Faculdade de Medicina, Universidade Eduardo Mondlane, Maputo, Mozambique; Department of Global Health, University of Washington, Seattle, Washington, USA; London School of Hygiene and Tropical Medicine, London, UK; Fundação Ariel Glaser Contra o SIDA Pediatrico, Maputo, Mozambique; Division of Global HIV and Tuberculosis at Centers for Disease Control and Prevention (CDC), Maputo, Mozambique; U.S Agency for International Development (USAID), Global Health, Washington, USA; National STI-HIV/AIDS Programme, Ministry of Health, Maputo, Mozambique

**Keywords:** HIV testing yield, Targeted provider-initiated HIV testing and counselling, Healthcare providers’ training, HIV positivity test, Mozambique, Implementation science

## Abstract

**Background:** In Mozambique, targeted provider-initiated HIV testing and counselling (PITC) is recommended to increase HIV testing yield where universal PITC is not feasible. However, its effectiveness depends on healthcare providers’ training. We evaluated the effect of a Ministry of Health training module in targeted PITC on the HIV testing yield.

**Methods:** We conducted a single-group pre-post study between November 2018 and November 2019 in the triage and emergency departments of four healthcare facilities in Manhiça District. It consisted of two two-month phases split by a one-week targeted PITC training module (“observation phases”). During both phases, providers reported their recommendation to test or not for individuals ≥15 years, and study HIV counsellors performed universal testing. We calculated HIV testing yield of targeted PITC as the proportion of HIV-positive individuals among those provider-recommended and tested. We compared pre- and post-training yields using two-proportion z-test. Additionally, we extracted aggregated health information system data over the four months preceding and following the observation phases to compare yield in real-world conditions (“routine phases”). We used logistic regression to identify predictors of HIV test positivity.

**Results:** Among 7,102 participants in the pre- and post-training observation phases (58.5% and 41.5% respectively), 68% were women, and 96% were recruited at triage. While HIV testing yield between pre- and post-training observation phases was similar, we observed an increase in yield in the post-training routine phase for women in triage (Yield ratio=1.54; 95%CI: 1.11-2.14). Age (25-49 years) (OR=2.43; 95%CI: 1.37-4.33), working in industry/mining (OR=4.94; 95%CI: 2.17-11.23), unawareness of partner’s HIV status (OR=2.50; 95%CI: 1.91-3.27), and visiting a healer (OR=1.74; 95%CI: 1.03-2.93) were factors associated with HIV test positivity. Including these factors in the targeted PITC algorithm could increase new HIV diagnoses by 2.6%. Furthermore, testing individuals with ≥1 HIV risk factor/symptom and a negative HIV test within the past three months revealed an additional 3.5% of undiagnosed PLHIV.

**Conclusions:** We found over 50% increase in the HIV testing yield of targeted PITC among women in the four months following the training and observation phases. Including additional sociodemographic and risk factors in the targeted PITC algorithm could help identify undiagnosed PLHIV.

## Background

The Joint United Nations Programme on HIV/AIDS (UNAIDS) set the “95-95-95” targets as part of the strategy to end the AIDS epidemic by 2030 [1]. These targets aim to ensure that by 2025, 95% of people living with HIV (PLHIV) know their HIV status, 95% of people diagnosed with HIV receive sustained antiretroviral therapy (ART), and 95% of those on ART achieve viral load suppression. The first 95 target is the single most important step as it enables access to treatment and care. In 2021, Eastern and Southern Africa remained the regions with the highest number of PLHIV, with 20.6 million ─54% of all PLHIV in the world [2]. While many countries of the region have achieved the second and third “95-95-95” targets, they still struggle with the first one [2]. By the end of 2021, the gap to achieve the first 95 was 1.3 million among all PLHIV in Eastern and Southern Africa. In Mozambique, one of the most severely affected countries in the world by the HIV epidemic, out of the 2.1 million PLHIV in 2021, 84% knew their status [3]. To address this first 95 target, the World Health Organization (WHO) recommends universal provider-initiated testing and counselling (PITC) for all individuals attending healthcare facilities in high HIV burden countries [4]. HIV testing yield, defined as the proportion of individuals who newly test positive for HIV out of the total individuals tested, has been often used to monitor the performance of HIV testing programmes [5,6].

As countries approach the “95-95-95” targets, the number of undiagnosed PLHIV decreases, leading to fewer new HIV infections. However, this also means that more HIV tests are needed to detect the remaining undiagnosed PLHIV, resulting in a decline in HIV testing yield. [7]. This along with limited resources and capacity has led to consideration of alternative HIV testing and counselling (HTC) modalities focused on people at higher risk [8].Targeted PITC, through which healthcare providers offer HTC to individuals presenting with risk factors, signs or symptoms suggestive of underlying HIV infection, has been shown to improve HIV testing yield and efficiency in many Sub-Saharan African (SSA) countries [9–11]. In Mozambique, targeted PITC is the modality that identifies the largest number of new HIV diagnoses, especially among adults over 25 years, with the national HIV testing yield in 2021 estimated at 6% [3,12]. Prioritization for HIV testing through targeted PITC depends on healthcare provider experience, training, daily patient load, and worker’s turnover. To optimize targeted PITC, the Mozambican Ministry of Health (MoH) developed a training module for healthcare providers [13].

Our objective was to evaluate the effect of a MoH training module in targeted PITC on the HIV testing yield in the triage and emergency department (ED) in southern Mozambique. We also aimed to identify further predictors of HIV test positivity beyond those already included in the existing national targeted PITC algorithm.

## Methods

### Study design, setting and population

We conducted a single-group pre-post study involving healthcare providers between November 2018 and November 2019 in the four highest-volume healthcare facilities of Manhiça District (MD) ─MD hospital, Xinavane rural hospital, and Palmeira and Maragra health units. MD is a semi-rural area in the Maputo province, southern Mozambique, with an overall community HIV-prevalence of 36.6% in 2015 [14,15]. The Mozambican national HIV testing strategy recommends targeted PITC in the high-volume entry points of healthcare facilities where universal PITC is not feasible to be applied, such as the triage and ED [16,17] [see Additional file 1].

The study consisted of two two-month individual-level data phases (“observation phases”) split by a one-week targeted PITC training module: i) pre-training observation phase (1 March-12 May 2019), ii) training (13-19 May 2019), iii) post-training observation phase (20 May-31 July 2019) (Figure 1). We invited healthcare providers who worked routinely in the triage and ED to participate, and we include those who agreed to participate in both study observation phases and in the targeted PITC training module. Inclusion criteria for clients included being ≥15 years old, resident of MD, presenting to the triage or ED with non-disabling condition for participating in the study and giving informed consent (IC). We did not include those individuals who were <18 years old and came to the healthcare facility without a caregiver. Additionally, in order to control for the potential observer bias effect and estimate the yield in real-world conditions, we extracted data from the health information data system of all individuals presenting at triage and ED four months preceding and following both observation phases and training (“routine phases”): iv) pre-training routine phase (1 November 2018-28 February 2019), v) post-training routine phase (1 August-30 November 2019) (Figure 1). These data were aggregated by healthcare facility, department, day, sex and age group.

**Figure 1.**
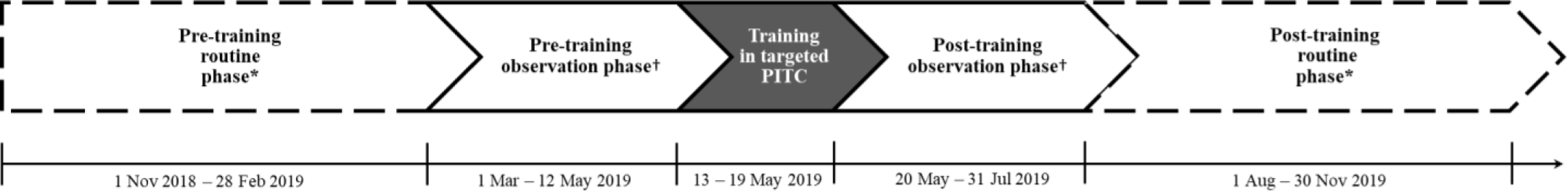
Study timeline. *Aggregated data extracted from the health information data system (real-world conditions). † Individual-level data collected through a study case report form (study conditions). <u>Abbreviations:</u> PITC: provider-initiated testing and counselling

### Targeted PITC algorithm and training module

The Mozambican national strategy for targeted PITC includes an algorithm based on risk factors, signs and symptoms to prioritize those individuals at higher risk for HIV testing [see Additional file 2] [16]. An individual is eligible for HIV testing through targeted PITC when he/she reports at least one of the symptoms or risk factors included in the algorithm, does not have a previous HIV diagnosis and does not have a negative HIV test result in the previous three months.

The training module in targeted PITC, based on the algorithm, consisted of one week of training aimed at optimizing healthcare providers’ prioritization for HIV testing through targeted PITC. It was designed and provided by the HTC reference group of the Mozambican MoH to ensure that it was standardised and reproducible [13]. At the time of the study, the training module was in a pilot phase, and it was on the verge of being implemented. Since then it has become a standardised MoH-approved training module, but its routine implementation is not mandatory.

### Study visit procedures for observation phases

During the routine clinic visit, healthcare providers recorded their decision and reason to refer or not refer individuals for HIV testing in a paper-based case report form. Following the routine clinic visit with the provider, a study HIV counsellor invited the individuals who met the inclusion criteria to participate in the study. After obtaining IC, we offered HIV testing to the participant regardless of the provider’s recommendation in order to assess the number of new HIV diagnosis missed by targeted PITC [see Additional file 1]. We did not test those participants with a previously known HIV diagnosis. The study HIV counsellor also collected clinical information (risk factors, signs and symptoms of HIV infection according to the national targeted PITC algorithm) and sociodemographic characteristics through an electronic study questionnaire. We included additional factors that have been described in the literature to be associated with HIV test positivity [9]. We performed the same study visit procedures in both observation phases.

### Statistical analysis

We calculated proportions for categorical variables and the median and interquartile range (IQR) for continuous variables. Categorial variables were compared using Pearson’s chi-squared test. We estimated HIV testing yield of targeted PITC as the proportion of HIV-positive individuals out of the total individuals referred by the provider and tested. We used the two-proportion z-test to test for the absolute difference in yield proportions of targeted PITC between the pre- and post-training observation phases and between the pre- and post-training routine phases, stratified by sex and health department. Moreover, we estimated yield ratios (YR) as a relative measure of associated gain using log-binomial regression models. We performed logistic regression analysis using individual-level data from the observation phases to assess the capacity of the signs and symptoms, risk factors and sociodemographic characteristics to predict HIV test positivity. We built a multivariable model by including all variables with a p-value <0.20 in bivariable analyses (sex and age were fixed), followed by backward stepwise selection, removing variables with p-values >0.20 and adding those with p-value <0.05. We used Stata version 16 for the analyses [18].

## Results

### Study profile for observation phases

A total of 19 healthcare providers, all medical assistants, participated in the study (three and six in the triage and ED of the MD hospital, respectively; two and two in the triage and ED of the Xinavane rural hospital, respectively; and three in the triage department of each of the Palmeira and Maragra health units). Out of the 8,533 clients screened for the study, we enrolled 83.2% as study participants ─4,155 (58.5%) and 2,947 (41.5%) in the pre- and post-training observation phase, respectively─ hereinafter referred to as client participants. Among them, 43.5% (n=1,809) and 45.7% (n=1,347) were eligible for HIV testing referral through targeted PITC in the pre- and post-training observation phases, respectively (Figure 2).

**Figure 2.**
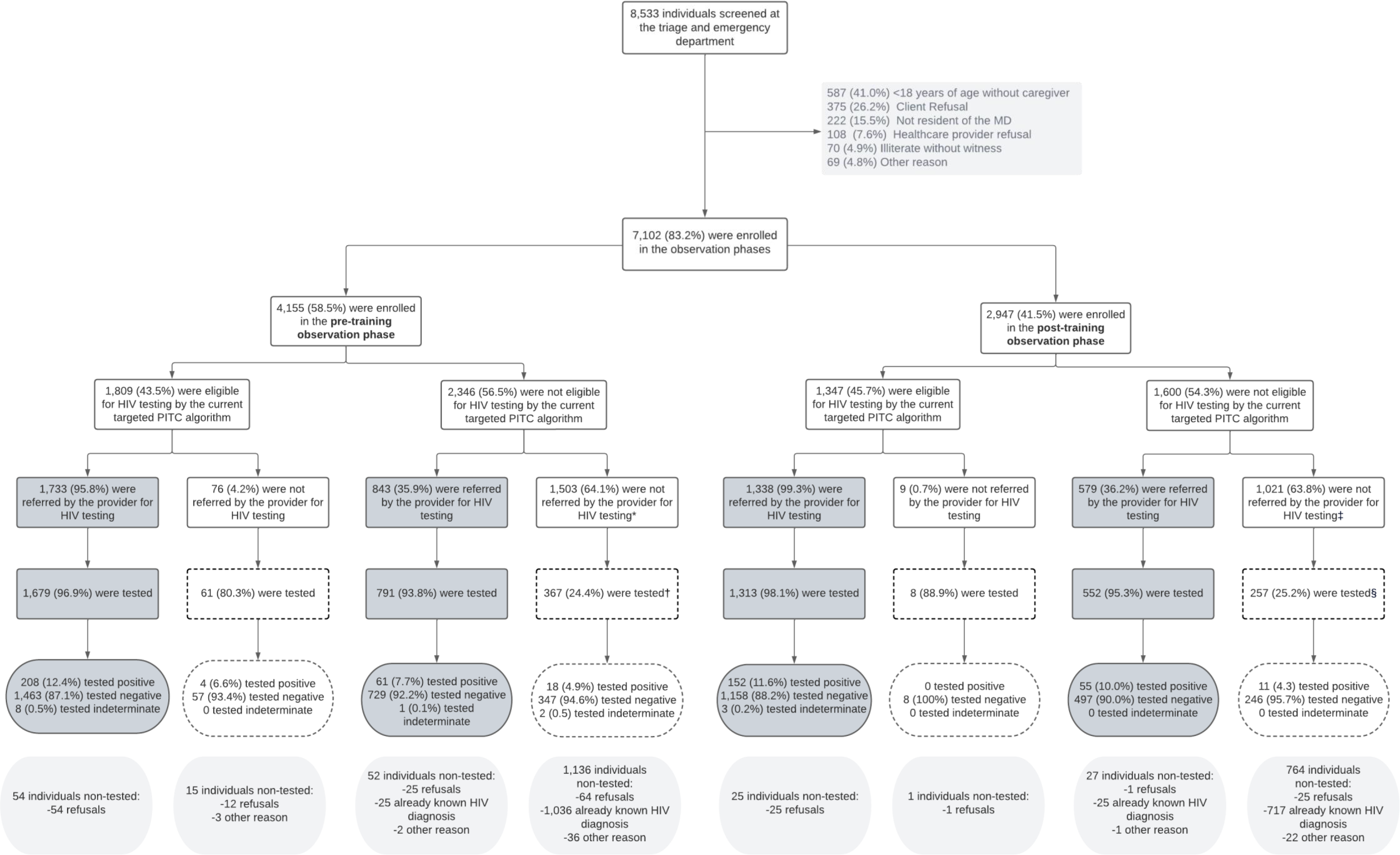
Study profile of the clients enrolled in the observation phases. * Including 384 individuals with a negative HIV test in the previous three months. † 367 individuals with a negative HIV test in the previous three months ‡ Including 269 individuals with a negative HIV test in the previous three months. § 257 individuals with a negative HIV test in the previous three months We defined eligible for HIV testing, according to the Mozambican national targeted PITC algorithm, as presenting with at least one HIV risk factor, sign or symptom and without an already known HIV diagnosis or negative HIV test in the previous three months. The areas shaded in dark grey correspond to the individuals tested by the targeted PITC modality for HIV testing and counselling. The boxes with dashed-lines correspond to individuals who were not tested by the targeted PITC modality, but who were tested because of study purposes. Study HIV counsellors tested all the individuals who did not have an already known HIV diagnosis. <u>Abbreviations:</u> PITC: provider-initiated testing and counselling

Healthcare providers referred 95.8% (1,733/1,809) of the algorithm-eligible individuals in the pre-training observation phase and 99.3% (1,338/1,347) in the post-training one. They also referred a third of algorithm-non-eligible individuals in the pre- and post-training observation phases: 35.9% (843/2,346) and 36.2% (579/1,600), respectively. Study HIV counsellors tested a total of 5,028 individuals without an already known HIV diagnosis, of whom 86.2% (n=4,335) were referrals from providers (Figure 2).

Providers stated two main reasons for not recommending for HIV testing: an already known HIV diagnosis (n=984; 62.3% and n=684; 66.4% in the pre- and post-training phases, respectively) and a negative HIV test in the previous three months (n=384; 24.3% and n=269; 26.1% in the pre- and post-training phases, respectively). Not presenting with HIV risk symptoms was only reported as a reason for not referring to HIV testing among 6.5% (n=101) and 1.1% (n=11) in both respective phases.

### Client characteristics

Most of the client participants were women (n=4,798; 68%), had a median age of 32 years (IQR: 23-45), and were recruited in the triage (n=6,792; 96%) and in the MD and Xinavane hospitals (n=2,565; 36% and n=2,110; 30%, respectively). Slightly more women than men (42.5% vs. 39.5%, p-value=0.015) and more individuals from the Xinavane rural hospital compared to the Maragra health unit (45.6% vs. 36.4, p-value<0.001) were recruited during the post-training observation phase (Table 1A).

**Table 1.**
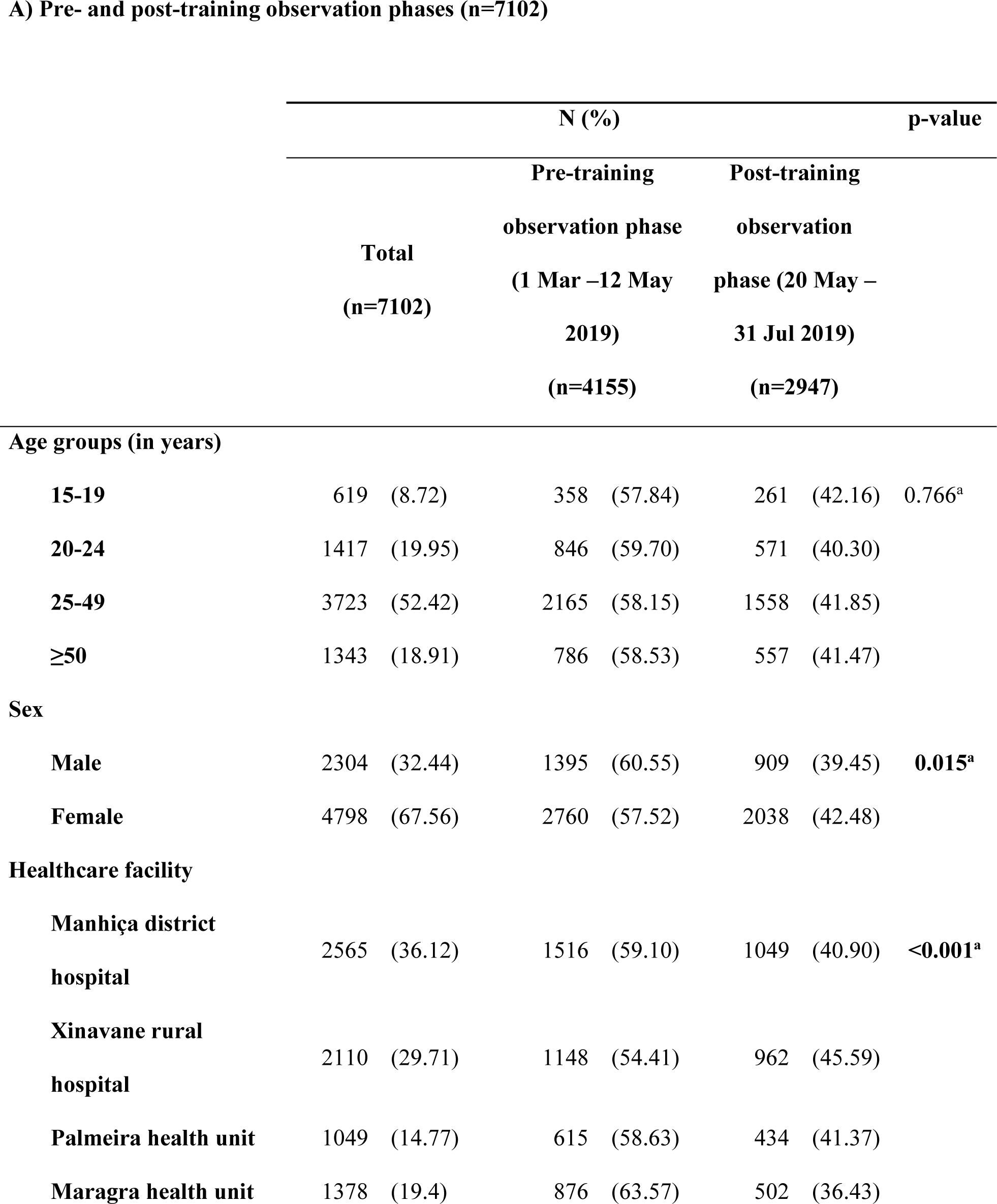
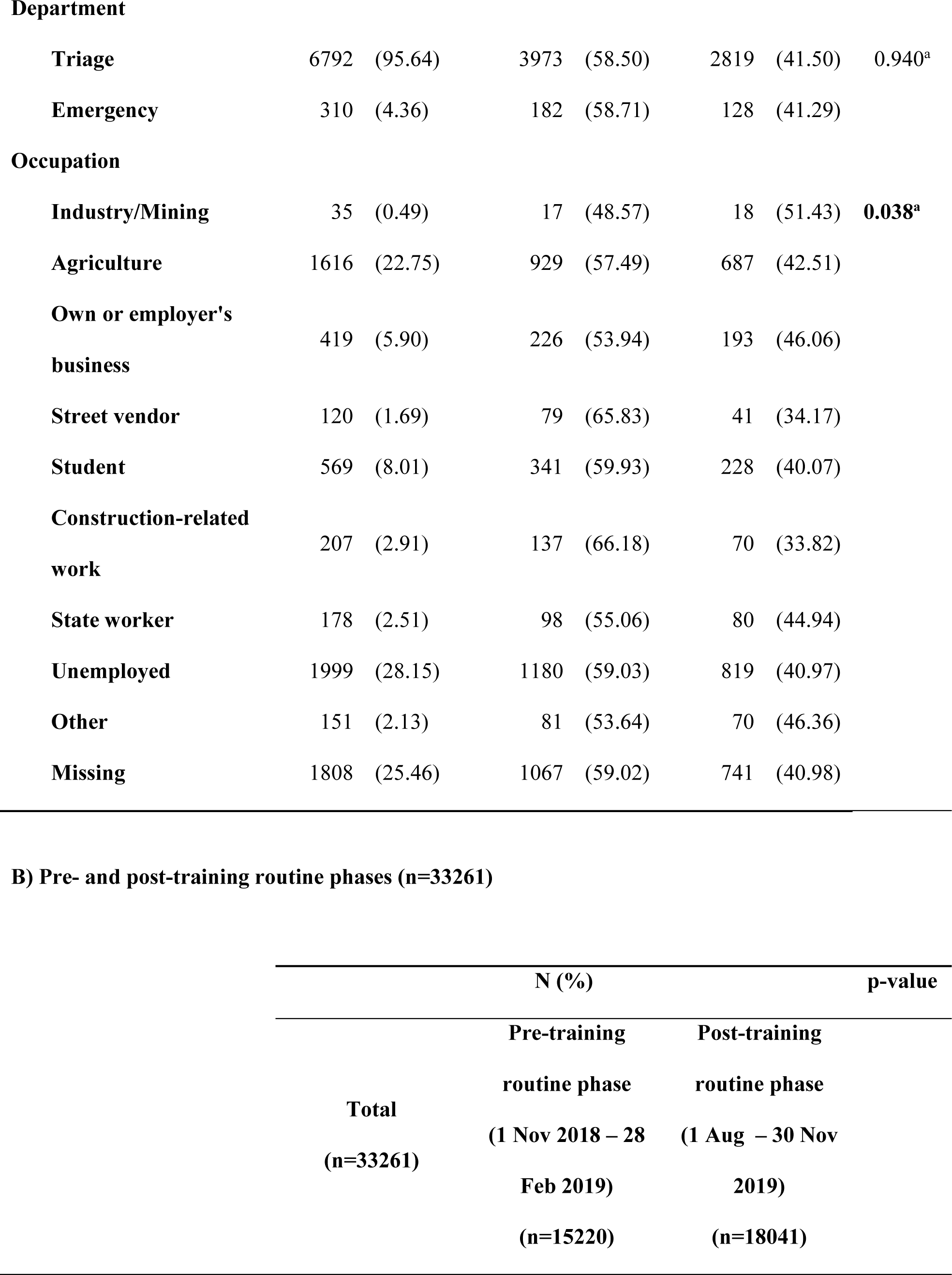
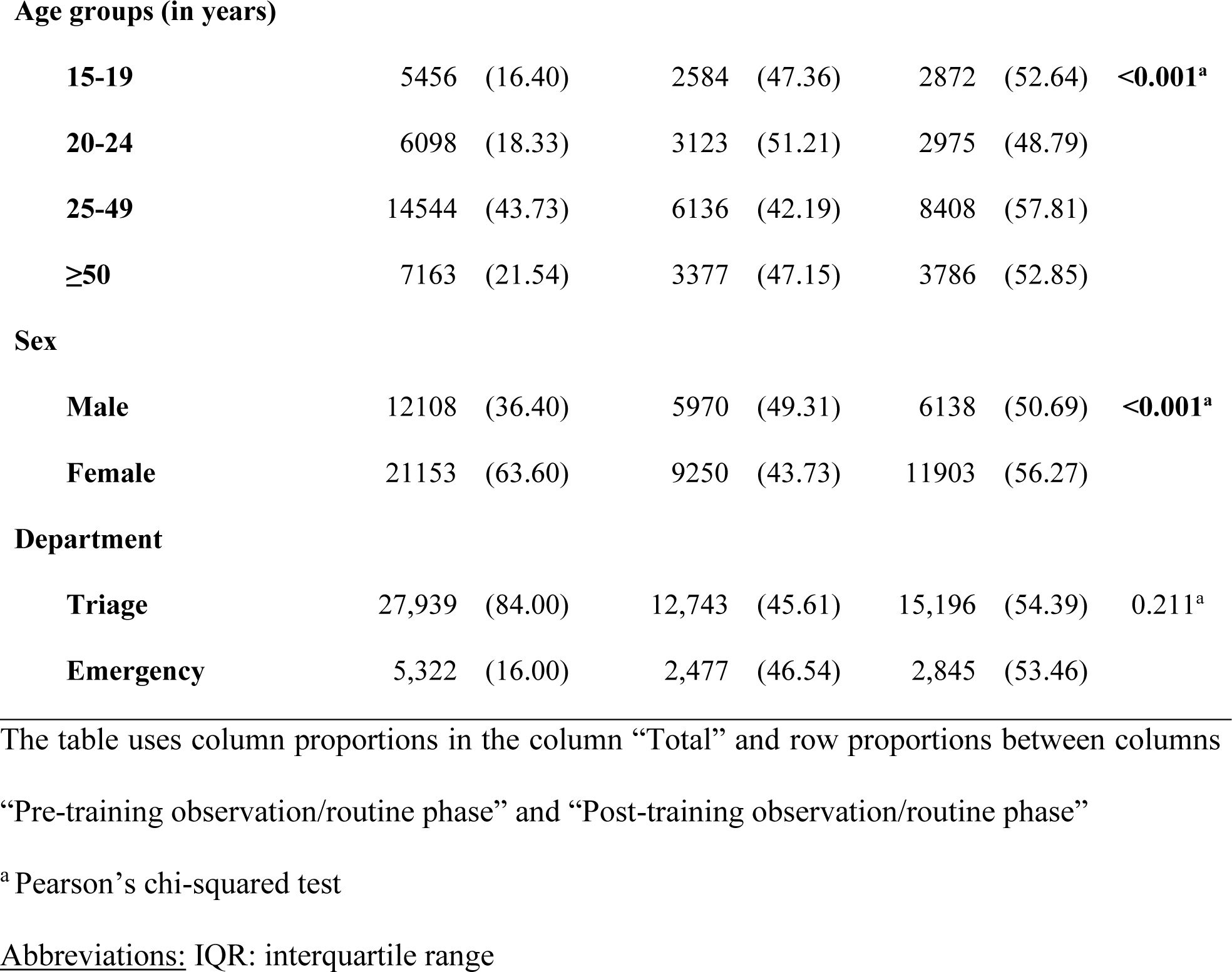
Client characteristics at enrolment.

When comparing characteristics from the pre- and post-training routine phases, we found that the proportion of women presenting at the healthcare facilities during the post-training phase was higher than men (56.3% vs. 50.7%, p<0.001), as well as, the proportion of individuals 25-49 years old compared to individuals 20-24 years old (57.8% vs. 48.8%; p<0.001) (Table 1B).

### HIV testing yield

The HIV testing yield of targeted PITC between the pre- and post-training observation phases was similar: 10.9% (95% confidence interval (CI): 9.7-12.2) and 11.1% (95%CI: 9.7-12.5), respectively (p-value=0.828). However, when we compared the yield between the pre- and post-training routine phases, the yield rose from 6.1% (95%CI: 5.2-7.0) to 8.9% (95%CI: 7.5-10.3) (p-value=0.001), representing a 46% increase in yield (YR=1.46; 95%CI: 1.18-1.81) (Table 2A).

**Table 2.**
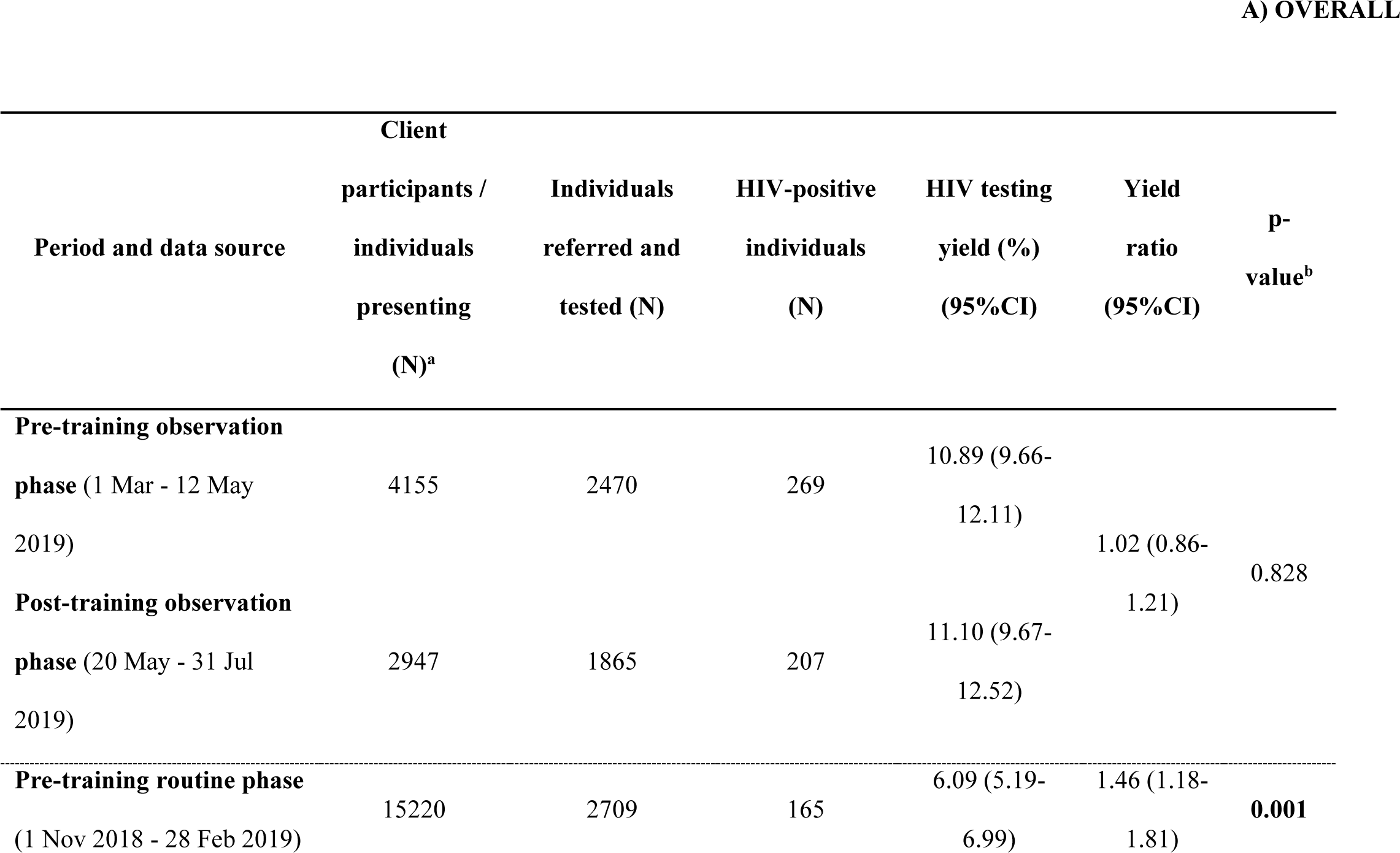
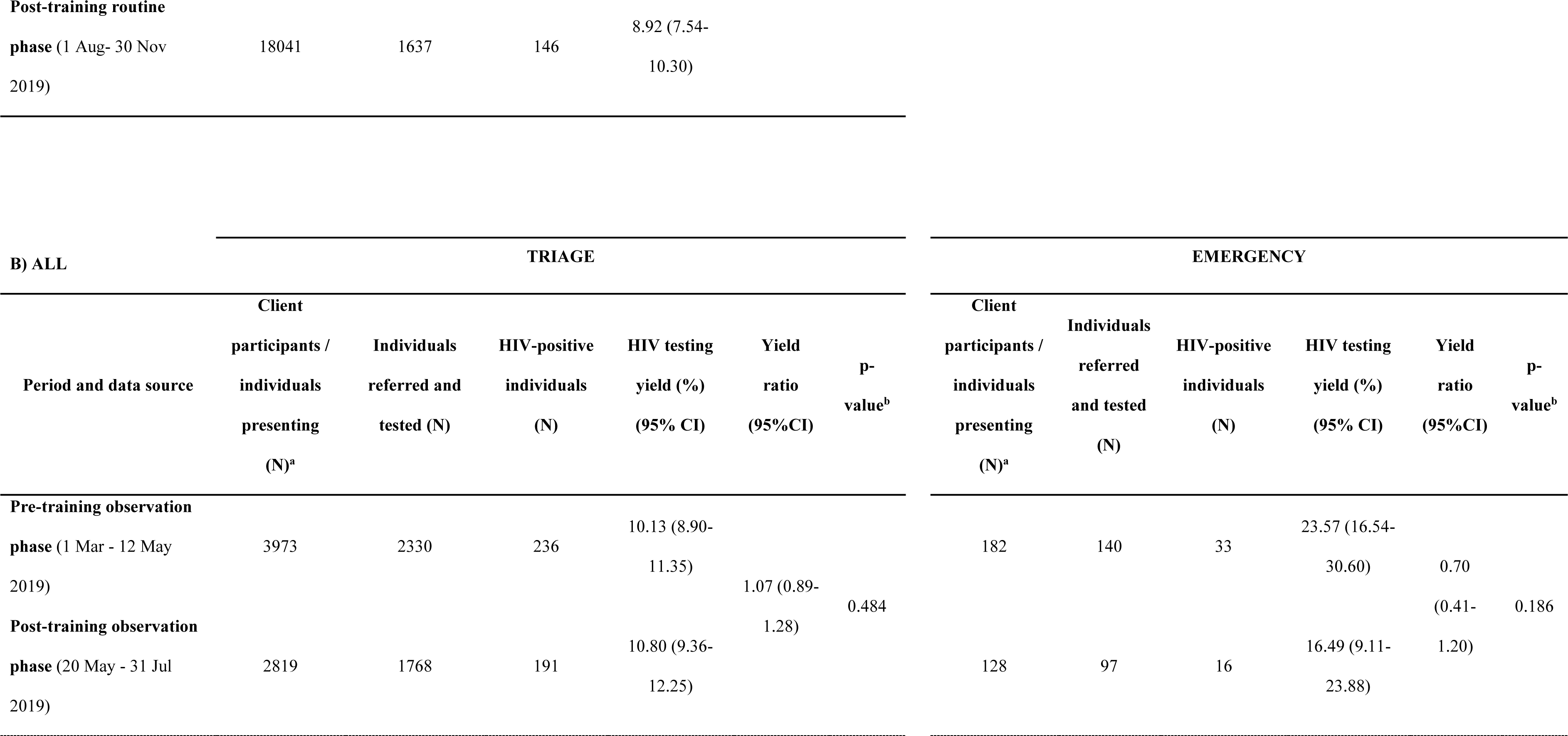
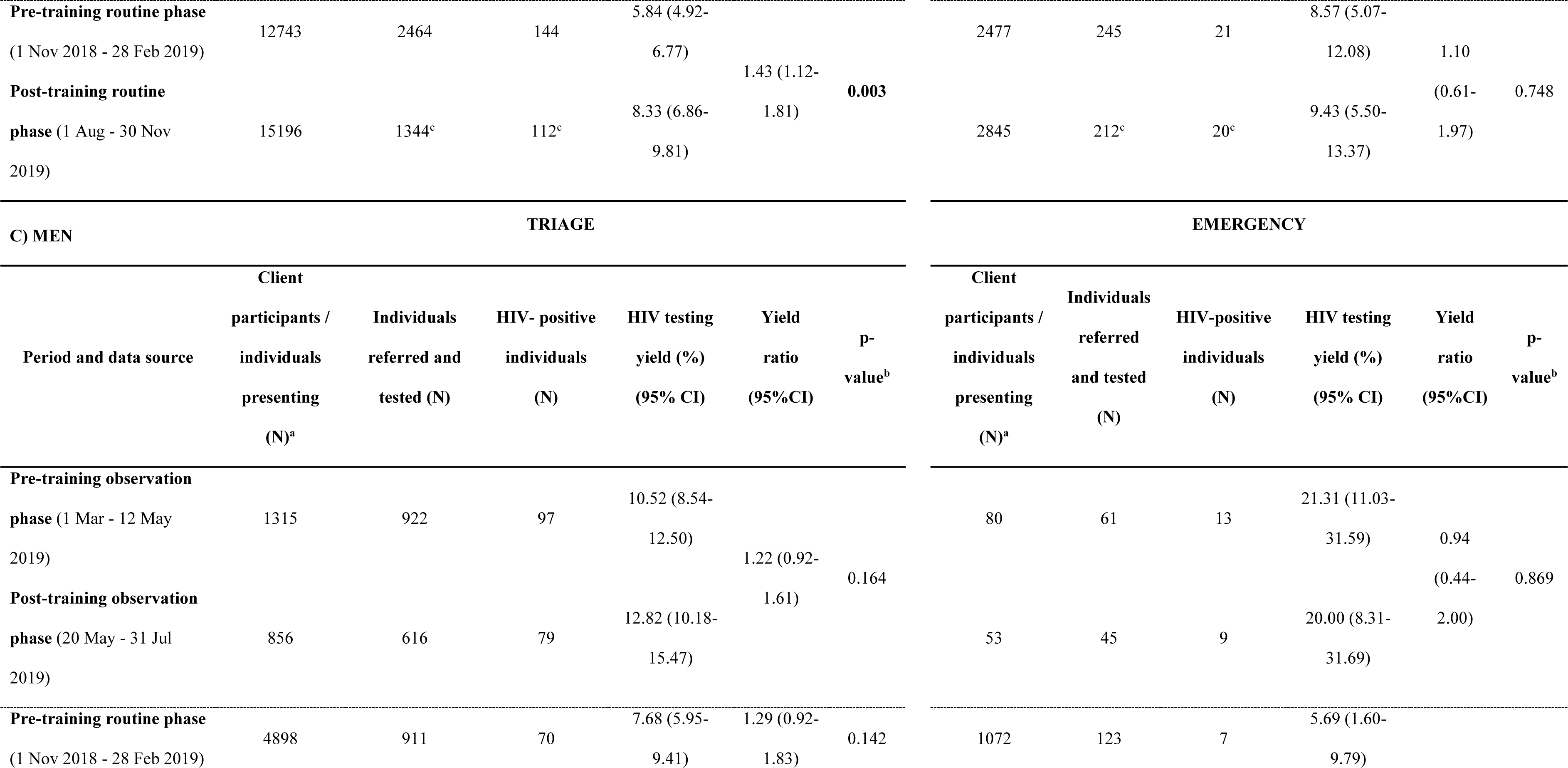
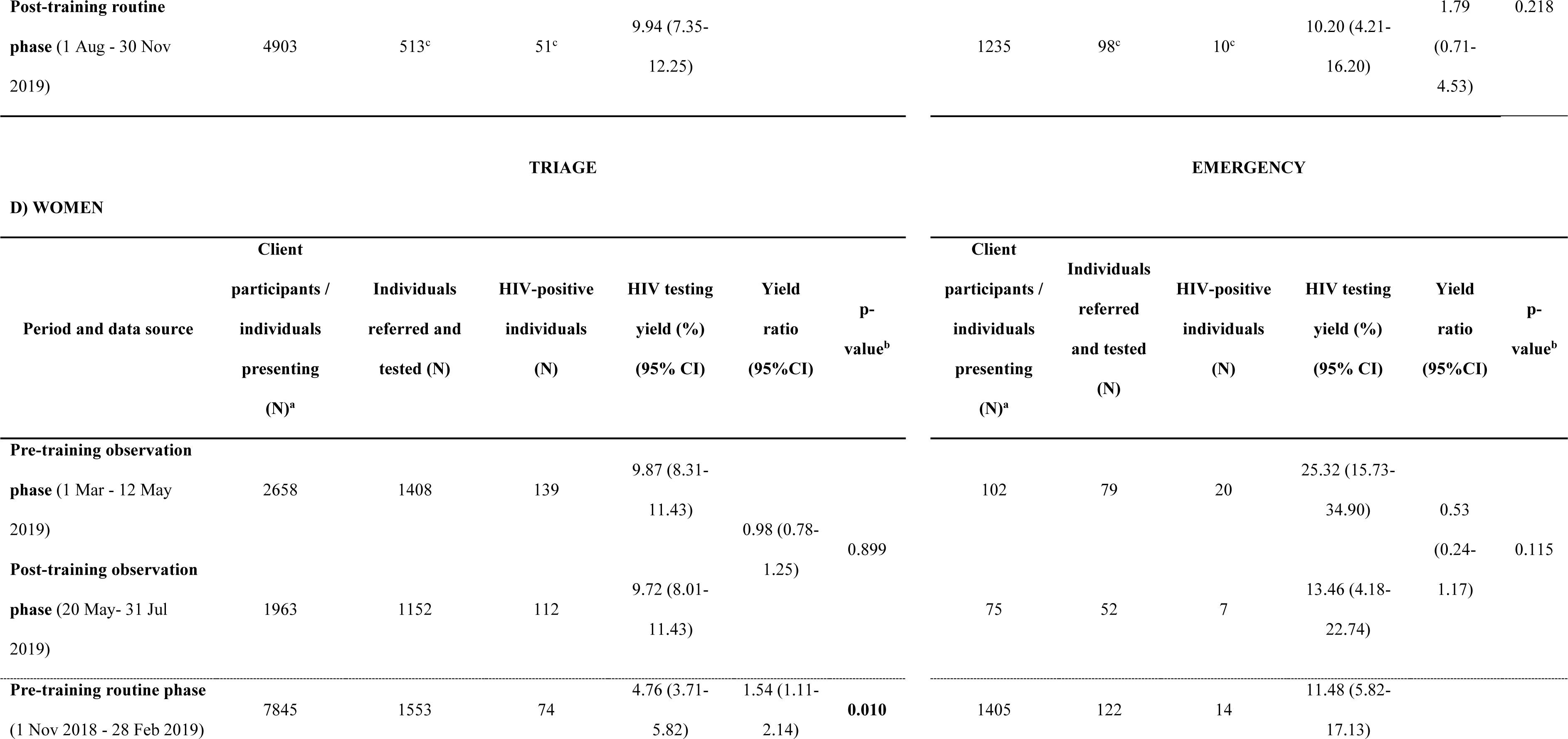
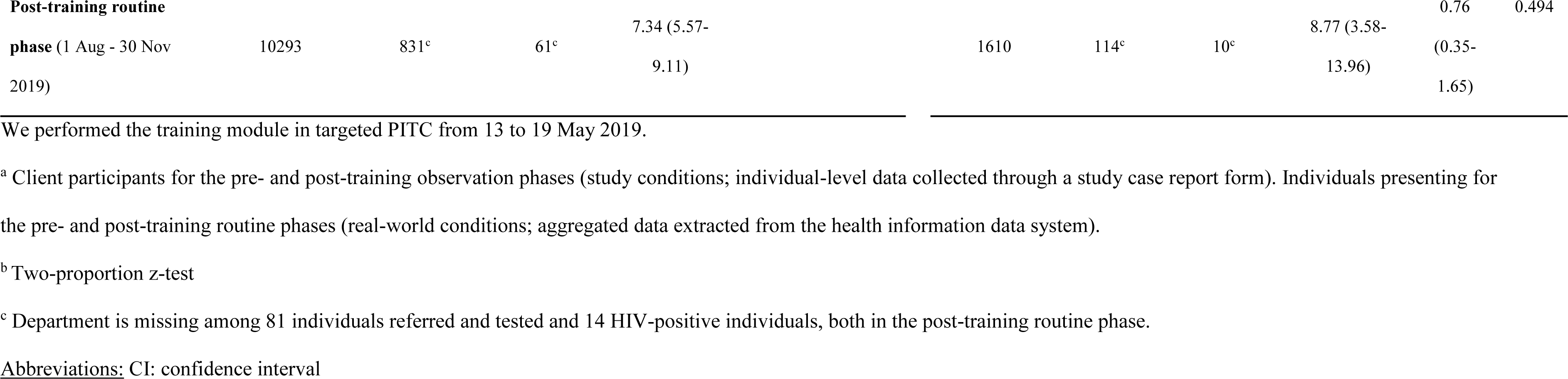
HIV testing yield of targeted PITC by study phase, health department and sex. Yield ratios from log-binomial regression model.

When stratifying by health department, the HIV testing yield in the triage rose from 5.8% (95%CI: 4.9-6.8) in the pre-training routine phase to 8.3% (95%CI: 6.9-9.8) in the post-training one (p-value=0.003). This represented a 43% increase in yield in triage in the months following the training and the observation phases (YR=1.43; 95%CI: 1.12-1.81). However, there was no significant increase in yield between the pre- and post-training routine phases in the ED (Table 2B).

Furthermore, when stratifying by sex and department, we found that the increase in yield in the triage during the post-training routine phase was significant among women (YR=1.54; 95%CI: 1.11-2.14) and not men (YR=1.29; 95%CI: 0.92-1.83). However, we observed greater yield values for men than for women in both the pre- and post-training routine phases (7.7%; 95%CI: 6.0-9.4 vs. 4.8%; 95%CI: 3.7-5.8, p-value=0.020 and 9.9%; 95%CI: 7.4-12.3 vs. 7.3%; 95%CI: 5.6-9.1, p-value=0.007, respectively) (Tables 2C and 2D).

### Predictors of HIV test positivity

We found a total of 509 (10.1%) individuals with a positive HIV test among the 5,028 individuals tested. HIV prevalence differed by sex (9.4% among women compared to 11.4% among men, p-value=0.025) and health department (9.7% in triage compared to 19.6% in the ED, p-value<0.001) (Table 3).

**Table 3.**
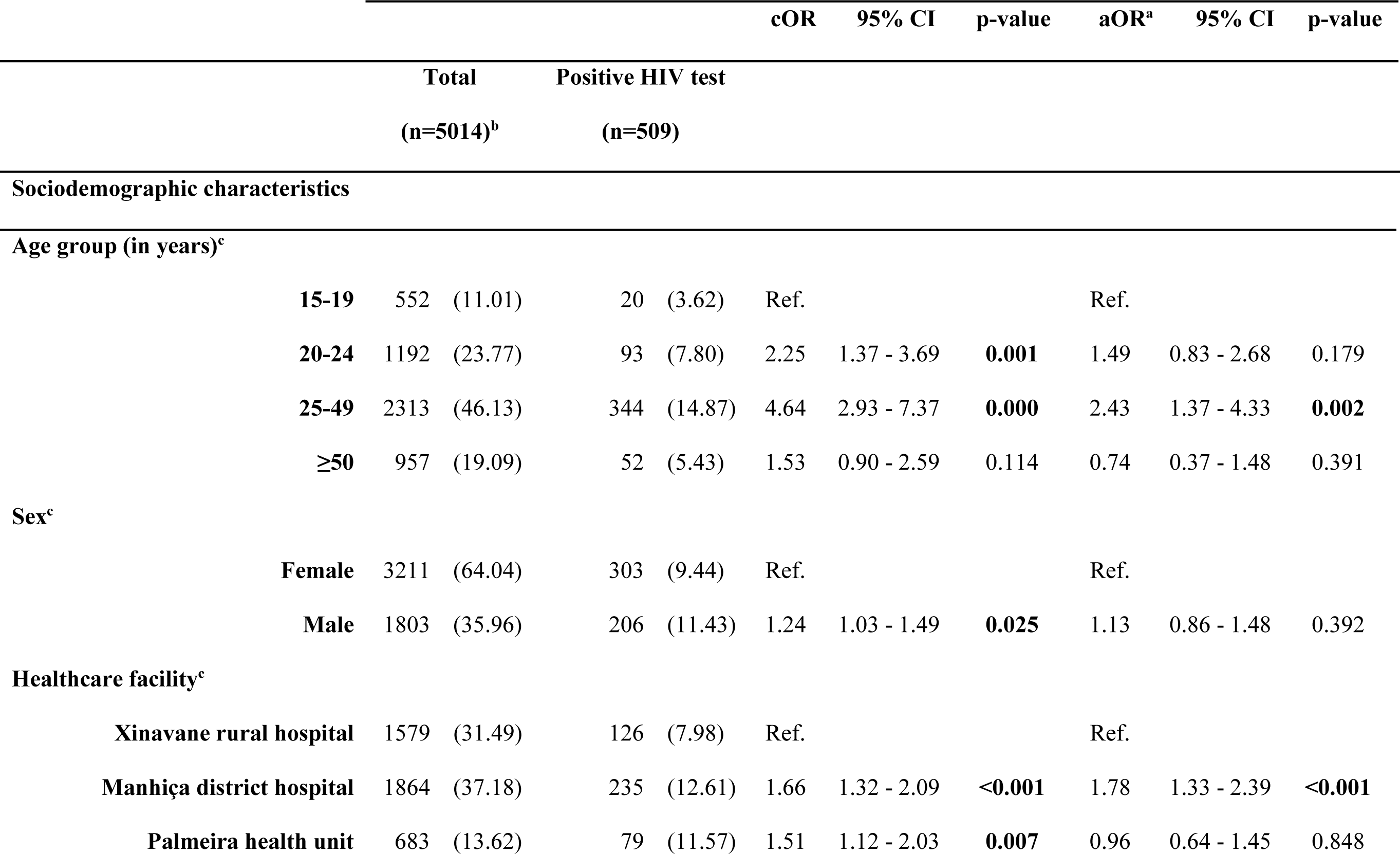
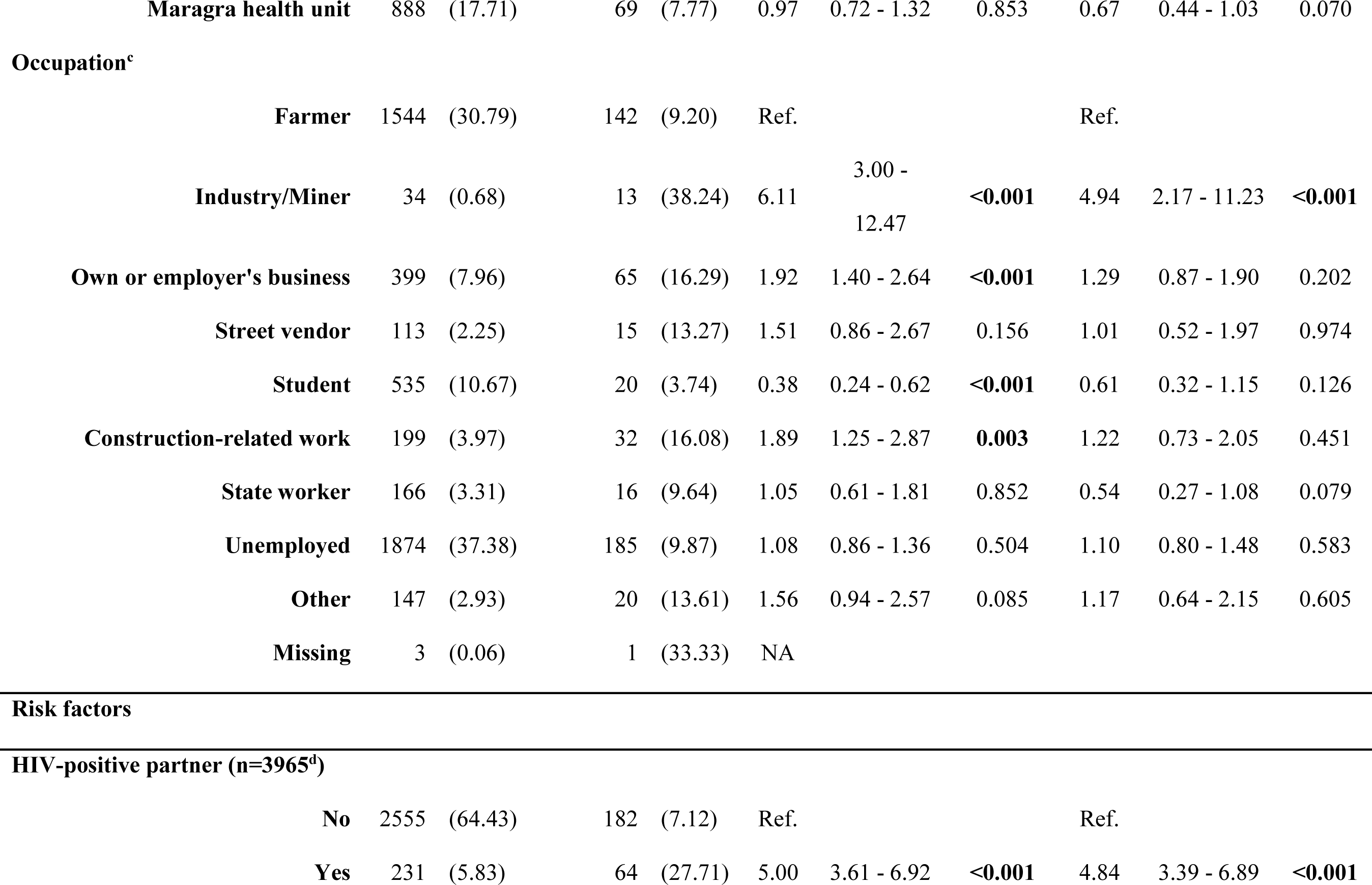
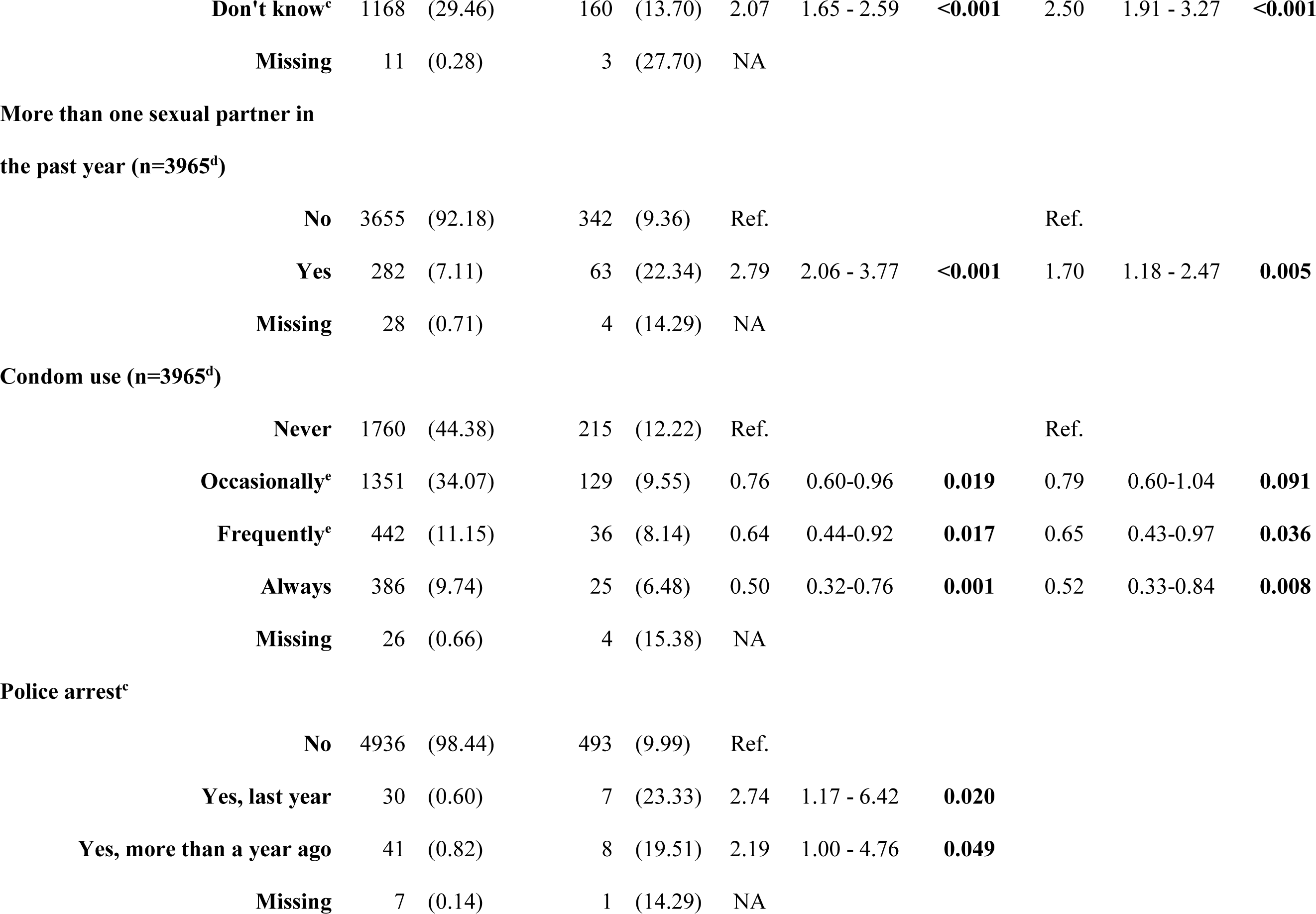
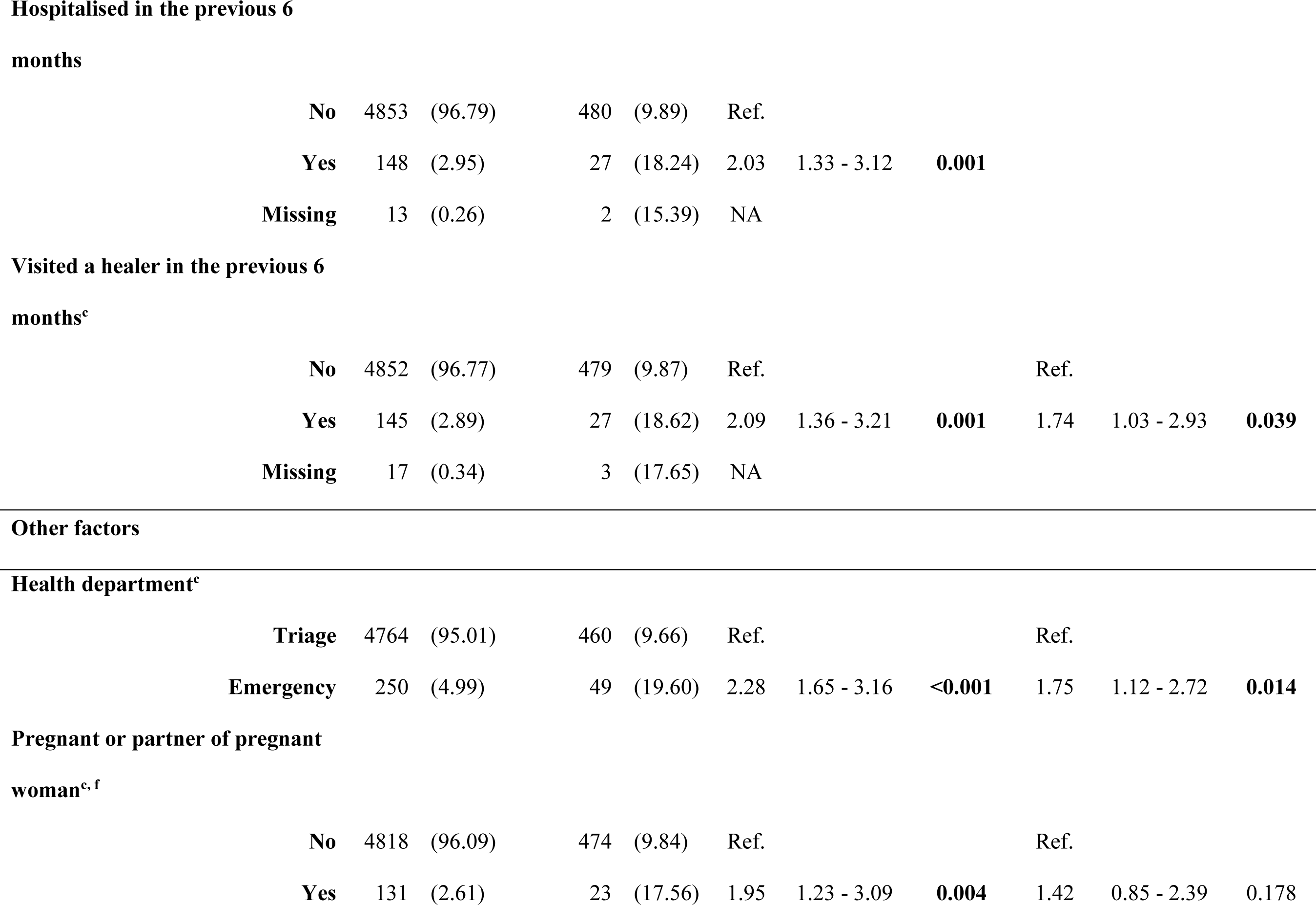
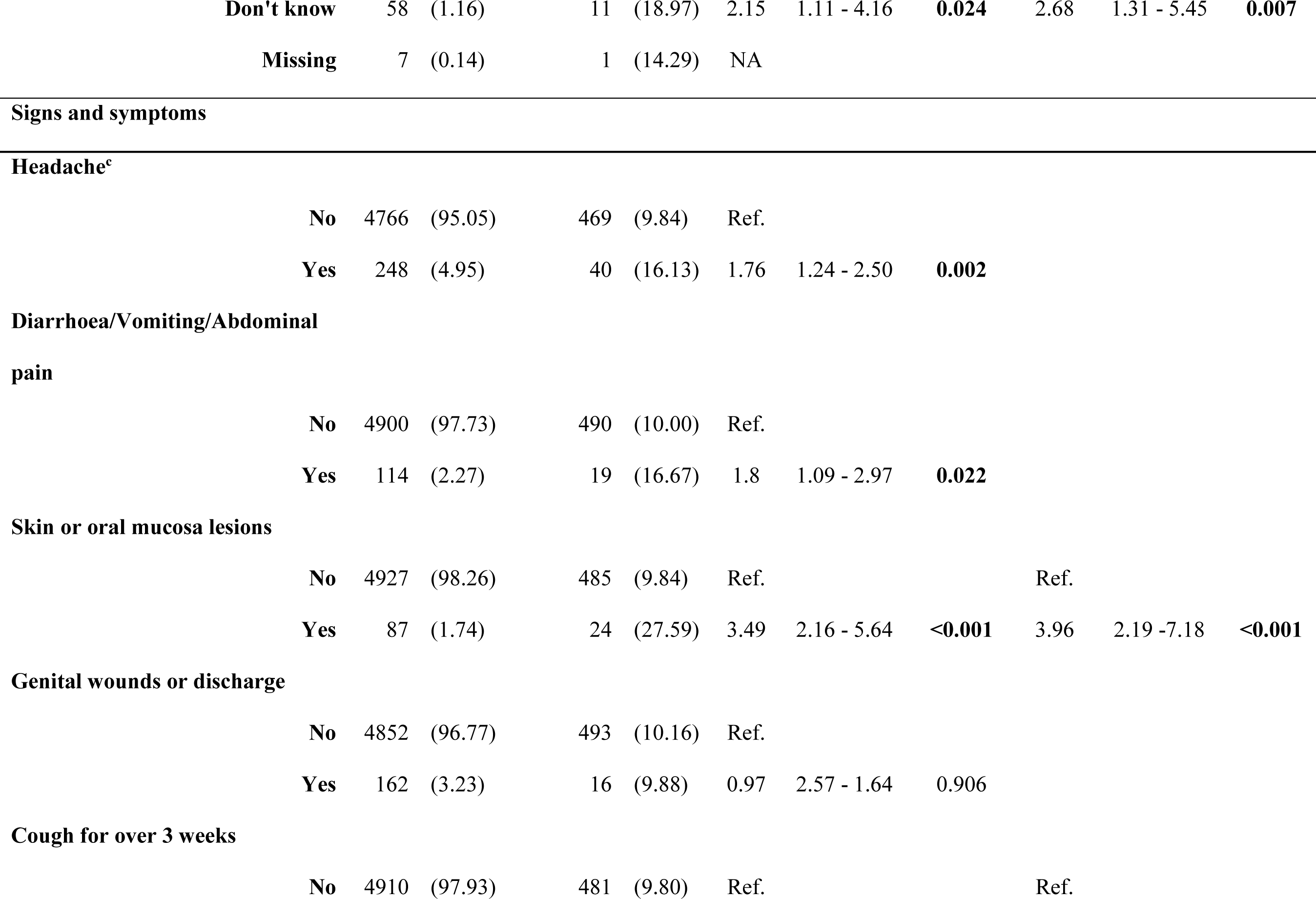
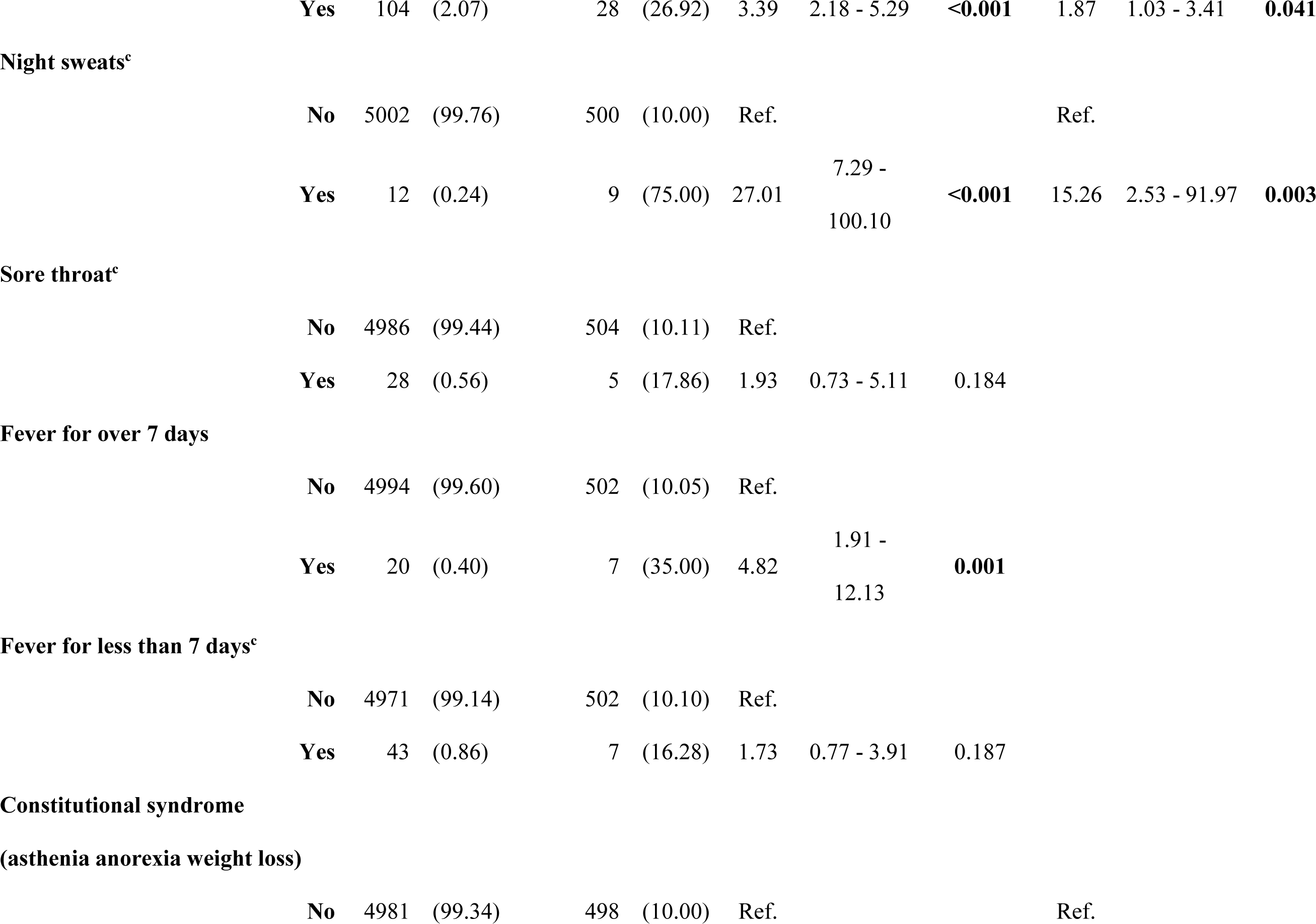
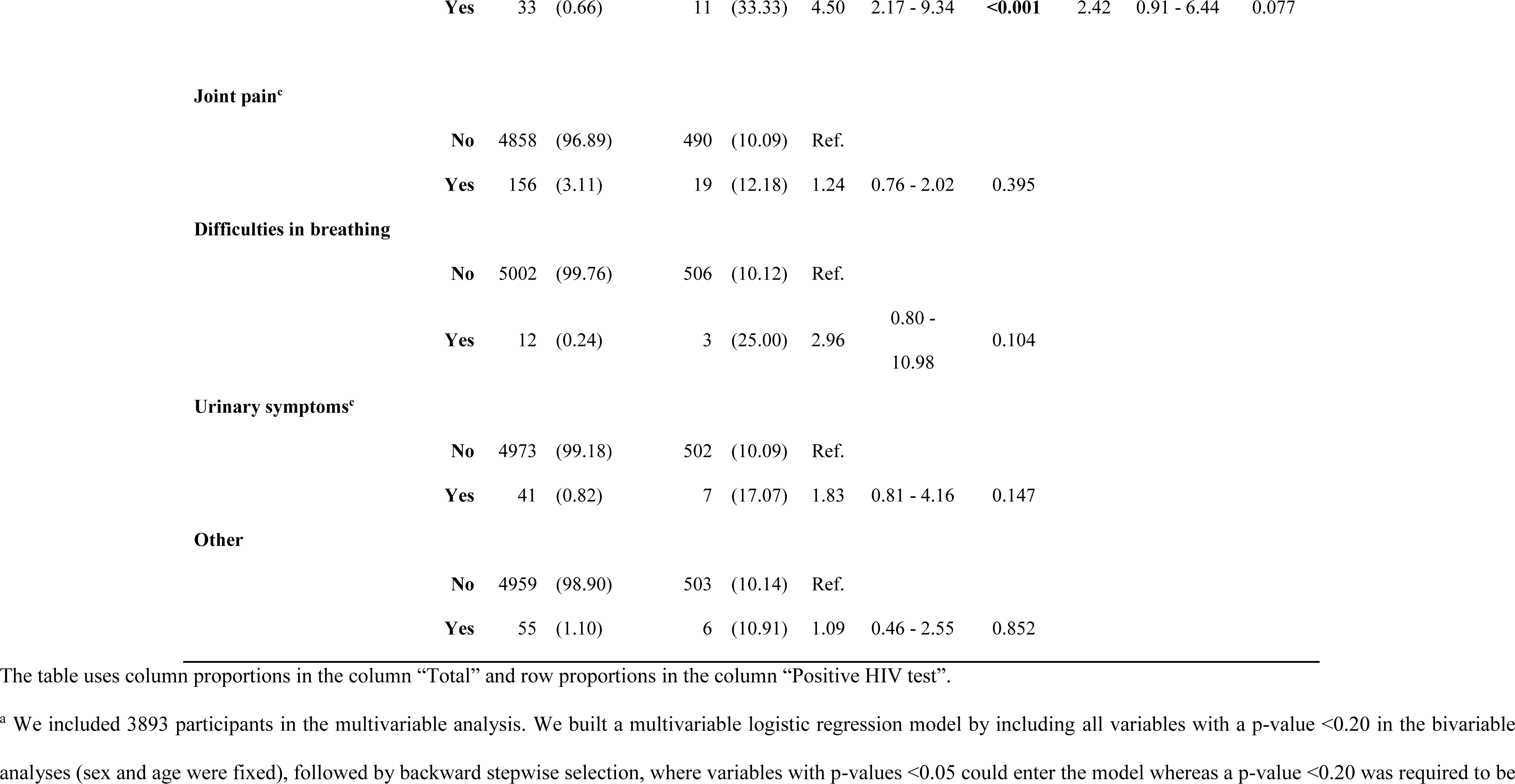
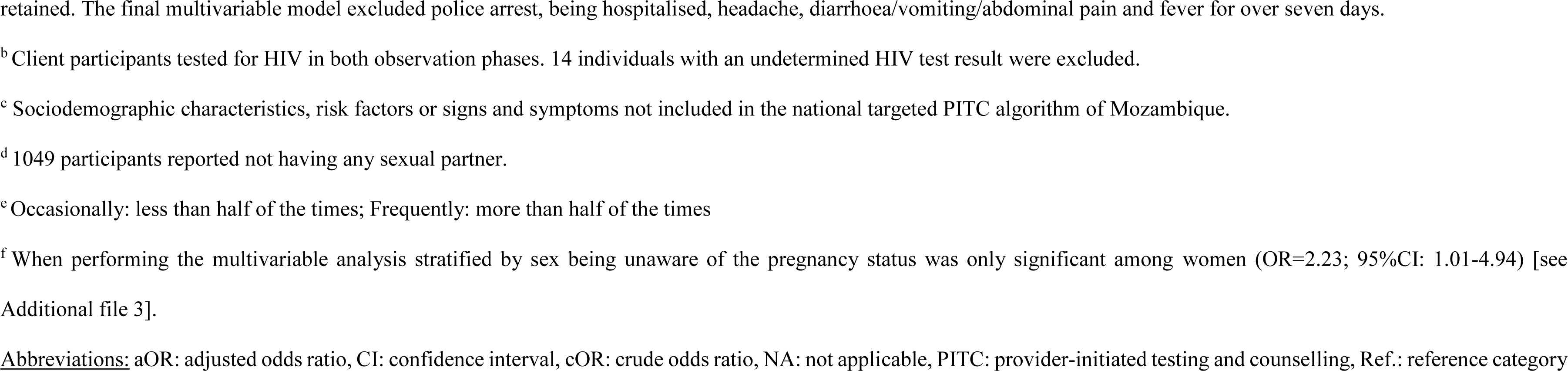
Factors associated with HIV test positivity. Crude and adjusted odds ratios from logistic regression analyses.

Multivariable analysis revealed that most of the variables included in the existing national targeted PITC algorithm were significantly associated with HIV test positivity. Among the other variables analysed, 25-49 years of age (OR=2.43; 95%CI: 1.37-4.33), presenting at the MD district hospital (OR=1.78; 95%CI: 1.33-2.39) and working in the industry or mining (OR=4.94; 95%CI: 2.17-11.23) were the sociodemographic factors associated with a positive HIV test. Regarding risk factors, unawareness of partner’s HIV status (OR=2.50 (95%CI; 1.91-3.27) and having visited a healer in the previous six months (OR=1.74; 95%CI: 1.03-2.93) were risk factors not included in the national PITC algorithm that showed a significant association with HIV test positivity. Likewise, presenting at the ED (OR=1.75; 95%CI: 1.12-2.72), unawareness of pregnancy status among women (OR=2.23; 95%CI: 1.01-4.94, [see Additional file 3]) and reporting night sweats (OR=15.26; 95%CI: 2.53-91.97) were other factors significantly associated with a positive HIV result (Table 3).

### Optimizing the targeted PITC algorithm

Out of the 509 individuals who tested positive for HIV, 72.0% (364/509) would have been identified by the existing national targeted PITC algorithm given a perfect interpretation. Regarding the factors not currently covered by the algorithm, there was no client unaware of the pregnancy status or who reported night sweats and who was not identified through the current algorithm as they presented at least one of the factors already included. In contrast, with the addition of age group, unawareness of partner’s HIV status, having visited a healer during the previous six months and working in industry or mining to the PITC algorithm, providers might have been able to reach 13 more undiagnosed PLHIV, which represents 2.6% (13/509) out of the individuals with a positive HIV test. Furthermore, testing individuals with at least one risk factor or symptom but a recent negative HIV test revealed 18 HIV-positive individuals. The current national targeted PITC algorithm excludes such individuals, but the removal of this criterion could increase new HIV diagnoses by 3.5% (18/509).

## Discussion

This study found that the HIV testing yield of targeted PITC increased by approximately 50% after the administration of a MoH training module to healthcare providers in the triage of four high-volume healthcare facilities in MD. This increase was observed in the post-training routine phase and specifically among women, while men had higher HIV testing yield values but showed no increase in the post-training phase. No differences were observed between the pre- and post-training observation phases. Additionally, we identified that age (25-49 years), occupation (industry or mining), unawareness of partner’s HIV status and a recent visit to a healer were factors associated with HIV test positivity that, if included in the targeted PITC algorithm, could increase the number of new HIV diagnoses by 2.6%. Furthermore, we found that the criterion of a negative HIV test in the previous three months, which excludes individuals from targeted PITC algorithm, leads to the missed opportunity of identifying an additional 3.5% of undiagnosed PLHIV.

During the observation phases, healthcare providers referred most of the targeted PITC algorithm-eligible individuals, but they also referred those individuals who did not present symptoms or risk factors for HIV and did not have an already known HIV diagnosis or a recent HIV negative test. The study itself is likely to have influenced providers to refer in a manner closer to universal PITC than to targeted PITC, even before they received training, due to their awareness of the ongoing evaluation of the PITC performance. This is known as the Hawthorne effect, which has been described in several SSA healthcare settings [19–22]. Additionally, resources such as the number of HIV counsellors were increased for study purposes, so providers knew that all individuals referred for testing during the observation phases would be tested, in contrast to routine conditions.

In real-world routine conditions, HIV testing yield among women presenting at triage was 54% higher in the post-training phase compared to the pre-training one. This suggests that the training module in targeted PITC may successfully increase HIV testing yield, as these differences were revealed when the study conditions ─Hawthorne effect and extra HIV counsellors─ were not present. Similar results were found in a study in Zambia, where providers trained in couples’ voluntary HTC increased their knowledge scores by 22% [23]. Furthermore, the fact that yield increased significantly among women, but not men, suggests that the training may be particularly effective in aiding diagnosis among a population with a low level of undiagnosed HIV. Conversely, men had greater HIV testing yield values than women in both pre- and post-training phases, which is consistent with findings from a study performed in Mozambique in 2019 [24] and could be explained by the higher prevalence of undiagnosed HIV among men (11.4% vs 9.4% in men and women respectively). Men are less likely to be tested and they have delayed entry to HIV care (first 95) in SSA countries like Mozambique [14,25], leading to a higher number of undiagnosed men living with HIV.

Of note, the current targeted PITC algorithm excludes individuals with a negative HIV test within three months prior. This exclusion criterion takes precedence over the inclusion criteria of HIV risk factors and symptoms. However, our results show that among these algorithm non-eligible individuals, some had at least one HIV risk factor or symptom included in the algorithm. The inclusion of these individuals for HIV testing could have identified an additional 3.5% of undiagnosed PLHIV. These new HIV diagnoses in this population reflect new infections within the three-month window and argue for the removal of this exclusion criterion from the algorithm in areas with high HIV incidence.

Presenting at the ED, a health department with a low volume of clients but a 20% prevalence of undiagnosed HIV, was associated with HIV test positivity. WHO guidelines recommend universal PITC in emergency services, and its implementation and enforcement could prevent missed opportunities for HIV diagnosis in Mozambique [26,27]. Studies in other countries, such as Tanzania, have shown that universal HIV testing in the ED is feasible with high acceptance and yield [28,29].

Moreover, the study found that most risk factors, signs and symptoms associated with HIV test positivity were previously described in the literature [9,30,31] and included in the Mozambican targeted PITC algorithm, which identified 72% of the positive HIV tests. Including additional variables such as age, occupation, unawareness of partner’s HIV status and recent visits to a healer could enhance the performance of the current algorithm, given their association with HIV test positivity, results consistent with previous studies [9,30,32,33]. Given complexities and time-pressures of including additional variables to the algorithm, the goal should first be to reinforce the existing algorithm.

In order to reach the UNAIDS first 95 target, it is widely accepted that as the pool of undiagnosed PLHIV is reduced, the number of tests to perform will increase [34]. Any changes to current algorithms should be evaluated in terms of cost-effectiveness, optimization of the HIV testing-decision making rule and competing needs to go the last mile in diagnosing PLHIV among key and hard-to-reach populations.

Very few studies have evaluated the effect of a targeted PITC training on HIV testing yield. To our knowledge, this is the first evaluation of a MoH training module performed in Mozambique. One major strength was the inclusion of pre- and post-training routine data analysis to estimate yield in real-world conditions. In contrast, there were several limitations due to the design of the study. The lack of a control group restricted the ability to draw causal inferences or account for secular trends [35]. Although the time interval between pre- and post- training was short, we were not able to infer whether the increase in yield seen in the post-training routine phase was primarily caused by the training module, or also by others factors that might be temporally coincident with it. Similarly, we could not control for fluctuations in the number of healthcare seekers or positive HIV tests observed in all phases, which could be due to seasonal patterns in clinical activities and health-seeking behaviour previously described in similar settings [36–38]. Additionally, due to the nature of health system aggregated data collection for the routine phases, paired data analysis was not possible, which may have hindered the control of differences between healthcare providers. More ideal study designs, such as cluster randomised trials or stepped-wedge at healthcare facility level in order to incorporate control facilities, were not feasible since the MoH training was about to be implemented and resources were limited.

## Conclusions

Targeted PITC’s performance, as measured by HIV testing yield, improved among women in a high-volume urgent care setting (triage) in the months following a MoH targeted PITC training study. Regular refresher trainings could enhance targeted PITC yield sustainably over time, and thus, further studies are needed to determine how to introduce the training, in terms of length and frequency. Additionally, to enhance the current targeted PITC algorithm, our findings suggest the need to consider removing the recent HIV-negative test criterion for those with HIV risk factors and adding factors such as age, occupation, unawareness of partner’s HIV status, and visits to a traditional healer. Adapting existing algorithms is crucial for reaching undiagnosed PLHIV, treating all PLHIV and eliminating HIV.

## Data Availability

The datasets generated and analysed during the current study are not publicly available due to privacy or ethical restrictions but are available from the corresponding author on reasonable request.

## List of abbreviations

AIDS: acquired immunodeficiency syndrome.
ART: antiretroviral therapy
CI: confidence interval
ED: emergency department
HIV: human immunodeficiency virus
HTC: HIV testing and counselling
IC: informed consent
IQR: interquartile range
IRB: Institutional review board
MD: Manhiça District
MoH: Ministry of Health
NA: not applicable
OR: odds ratio
PITC: provider-initiated testing and counselling
PLHIV: people living with human immunodeficiency virus
Ref.: reference category
SSA: Sub-Saharan African
UNAIDS: Joint United Nations Programme on HIV/AIDS
WHO: World Health Organization
YR: yield ratio

## Declarations

## Ethics approval and consent to participate

This study was approved by the Mozambican MoH Institutional review board (IRB) (Approval Nr. 363/CNBS/18), the Centro de Investigação em Saúde de Mahiça IRB (Approval Nr. CIBS-CISM/010/2018) and the Hospital Clinic Barcelona IRB (Approval Nr. HCB/2019/0379) and reviewed according to Centers for Disease Control and Prevention human research protection procedures (Approval Nr. 009F3EB81AA4251). All study participants, both healthcare providers and individuals presenting at the healthcare facilities, completed IC. For participants aged 15-18 years, a parent/legal representative’s additional consent was necessary.

## Consent for publication

Not applicable.

## Availability of data materials

The datasets generated and/or analysed during the current study are not publicly available due to privacy or ethical restrictions but are available from the corresponding author on reasonable request.

## Authors’ contributions

SFL, TN, LFS, ELV, EB, SW, PK and DN were responsible for conceptualization and study design. ASL, SFL, and TS, conducted the data analysis. ASL, SFL, TN, LFS, ELV, OA, TS, and DN interpreted the data. ASL, SFL, and TS, drafted the initial version of the paper. SFL, LFS, ELV, OA, PV, SW, PK, NH, PY, GA, FB, and DN critically reviewed the data and revised the manuscript. All authors read and approved the final manuscript.

## Acknowledgements

We acknowledge support from the grant CEX2018-000806-S funded by MCIN/AEI/ 10.13039/501100011033, and support from the Generalitat de Catalunya through the CERCA Programme. The authors gratefully acknowledge the staff at Centro de Investigação em Saúde de Manhiça, in the Manhiça District, Mozambique, who worked to collect and manage the data, the Ministry of Health of Mozambique, our research team, collaborators, and especially all communities and participants involved.

## Additional material

Additional file 1: Supplementary Material 1

Additional file 2: Supplementary Figure 1

Additional file 3: Supplementary Table 1

