## Additional file 1: Supplementary Material 1 for "Enhancing HIV testing yield in southern Mozambique: the effect of a Ministry of Health training module in targeted provider-initiated testing and counselling"

Study setting

MD is served by one referral district hospital, one rural hospital and 15 peripheral health units which offer free HIV services.

Universal PITC is implemented in: antenatal clinic, maternity, high-risk paediatric clinic, inpatient ward, tuberculosis services, pre-operatory patients, blood bank referrals, postpartum consultation, and circumcision clinic.

Triage is the first point of contact for clients who arrive at the health facility seeking medical attention. This department is responsible for rapidly assessing the severity of the client's condition and prioritizing them based on the urgency of their medical needs. If a client presents with severe or life-threatening conditions he/she is immediately referred to the ED.

Study procedures

HIV testing was conducted following the Mozambican HIV testing algorithm which included two serial rapid diagnostic tests, Determine and Unigold (1).

Supplementary References

1. Directriz nacional para a implementação do aconselhamento e testagem em saúde. Ministério da Saúde (MISAU), Direcção Nacional de Assistência Médica (DNAM) para Implementação do Aconselhamento e Testagem em Saúde.

Mozambique; 2015.
