## Additional File 2: Supplementary Figure 1 for "Enhancing HIV testing yield in southern Mozambique: the effect of a Ministry of Health training module in targeted provider-initiated testing and counselling"

**Supplementary Figure 1. Targeted PITC algorithm implemented by the Ministry of Health (MoH) in Mozambique.**

**SCREENING FOR RISK FACTORS, SIGNS AND SYMPTOMS OF HIV IN ADULTS**

**Risk factors:**

1. Have you been hospitalised during the previous 6 months?
2. Do you have a sexual partner living with HIV?
3. Do you have more than one sexual partner?
4. Do you use condoms during sex?

**Signs and symptoms:**

1. Persistent or repeated respiratory signs and symptoms
2. Genital discharge or sores
3. Swelling of the lower limbs or lymph nodes
4. Noticeable weight loss
5. Diarrhoea for more than 15 days
6. Persistent unexplained fever
7. Mouth sores or difficulty eating
8. Skin problems (itching, night burn, sores)

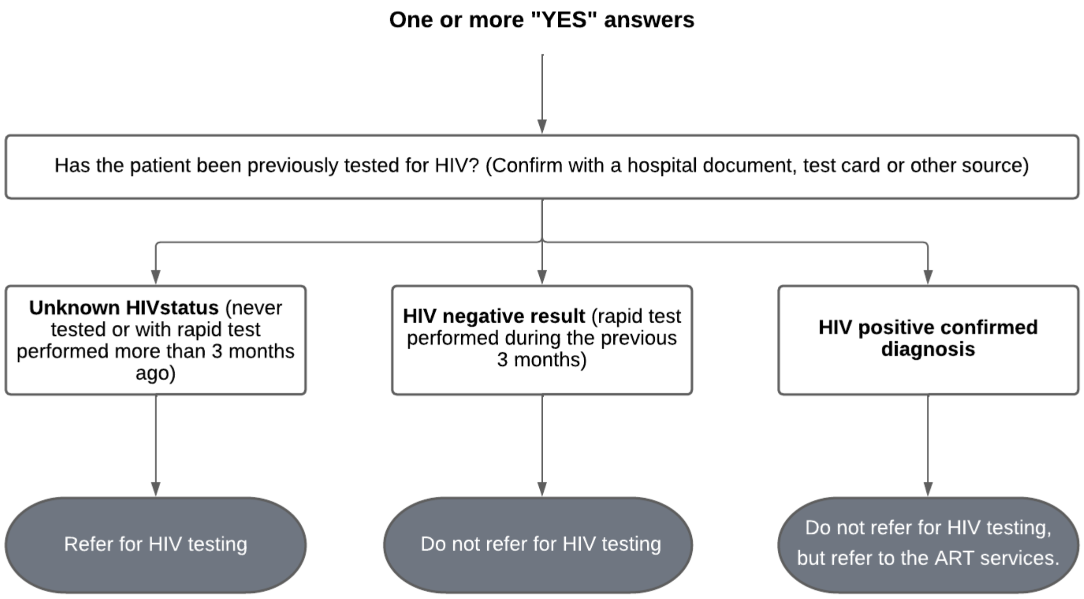


Figure adapted from the Differentiated Services Delivery Models Guidelines by the Mozambican MoH, 2018.

Abbreviations: ART: antiretroviral therapy, PITC: provider-initiated testing and counselling
