## Additional file 3: Supplementary Table 1 for "Enhancing HIV testing yield in southern Mozambique: the effect of a Ministry of Health training module in targeted provider-initiated testing and counselling"

**Supplementary Table 1. Factors associated with HIV test positivity. Adjusted odds ratios from logistic regression analyses by sex.**

| | **Men (n=1471)[a]** | | | **Women (n=2422)[a]** | | |
|---|---|---|---|---|---|---|
| | **aOR[a]** | **95% CI** | **p-value** | **aOR[a]** | **95% CI** | **p-value** |
| **Sociodemographic variables** | | | | | | |
| **Age group (in years)[b]** | | | | | | |
| **15-19** | Ref. | | | Ref. | | |
| **20-24** | 3.00 | 0.36-24.68 | 0.307 | 1.52 | 0.81-2.84 | 0.187 |
| **25-49** | 13.61 | 1.72-107.66 | **0.013** | 1.59 | 0.85-2.98 | 0.147 |
| **≥50** | 4.66 | 0.55-39.35 | 0.158 | 0.43 | 0.18-1.04 | 0.061 |
| **Health facility[b]** | | | | | | |
| **Manhiça district hospital** | Ref. | | | Ref. | | |
| **Xinavane rural hospital** | 0.82 | 0.52-1.30 | 0.404 | 0.41 | 0.27-0.61 | **<0.001** |
| **Palmeira health unit** | 0.64 | 0.33-1.25 | 0.191 | 0.52 | 0.32-0.83 | **0.007** |
| **Maragra health unit** | 0.51 | 0.26-1.02 | 0.057 | 0.27 | 0.16-0.48 | **<0.001** |
| **Occupation[b]** | | | | | | |
| **Farmer** | Ref. | | | Ref. | | |
| **Industry/Miner** | 3.81 | 1.55-9.34 | **0.003** | 7.66 | 0.62-94.05 | 0.112 |
| **Own or employer's business** | 0.98 | 0.57-1.67 | 0.935 | 1.75 | 0.98-3.11 | 0.057 |
| **Street vendor** | 0.42 | 0.52-3.47 | 0.424 | 1.35 | 0.64-2.84 | 0.425 |
| **Student** | 0.29 | 0.04-2.37 | 0.247 | 0.69 | 0.34-1.39 | 0.298 |
| **Construction-related work** | 1.03 | 0.57-1.84 | 0.928 | 5.10 | 0.91-28.47 | 0.063 |
| **State worker** | 0.52 | 0.23-1.18 | 0.117 | 0.42 | 0.10-1.82 | 0.247 |
| **Unemployed** | 0.91 | 0.54-1.55 | 0.729 | 1.29 | 0.86-1.92 | 0.217 |
| **Other** | 0.85 | 0.34-2.10 | 0.720 | 1.45 | 0.61-3.46 | 0.402 |

| | | | | | | |
|---|---|---|---|---|---|---|
| **Risk factors** | | | | | | |
| **HIV-positive partner** | | | | | | |
| **No** | Ref. | | | Ref. | | |
| **Yes** | 6.48 | 3.75-11.18 | **<0.001** | 4.12 | 2.52-6.74 | **<0.001** |
| **Don't know (The partner never told him/her)[b]** | 2.69 | 1.65-4.38 | **<0.001** | 2.80 | 2.00-3.92 | **<0.001** |
| **More than one sexual partner in the past year** | | | | | | |
| **No** | Ref. | | | Ref. | | |
| **Yes** | 1.65 | 1.04-2.64 | **0.034** | 2.21 | 1.10-4.43 | **0.026** |
| **Condom use** | | | | | | |
| **Always** | Ref. | | | Ref. | | |
| **Frequently[c]** | 1.81 | 0.78-4.21 | 0.171 | 1.27 | 0.55-2.95 | 0.577 |
| **Occasionally[c]** | 1.46 | 0.75-2.83 | 0.261 | 1.86 | 0.89-3.90 | 0.099 |
| **Never** | 1.77 | 0.92-3.42 | 0.089 | 2.30 | 1.11-4.79 | **0.026** |
| **Visited a healer in the previous 6 months[b]** | | | | | | |
| **No** | Ref. | | | Ref. | | |
| **Yes** | 2.10 | 0.95-4.63 | 0.066 | 1.59 | 0.76-3.31 | 0.217 |
| **Other factors** | | | | | | |
| **Health department [b]** | | | | | | |
| **Triage** | Ref. | | | Ref. | | |
| **Emergency** | 1.50 | 0.76-2.94 | 0.239 | 2.23 | 1.23-4.05 | **0.008** |
| **Pregnant or partner of pregnant woman[b]** | | | | | | |
| **No** | Ref. | | | Ref. | | |

| | | | | | | |
|---|---|---|---|---|---|---|
| **Yes** | 1.50 | 0.82-2.73 | 0.187 | 1.52 | 0.46-5.05 | 0.492 |
| **Don't know** | 2.82 | 0.50-15.90 | 0.239 | 2.23 | 1.01-4.94 | **0.048** |
| **Signs and symptoms** | | | | | | |
| **Skin or oral mucosa lesions** | | | | | | |
| **No** | Ref. | | | Ref. | | |
| **Yes** | 3.54 | 1.50-8.35 | **0.004** | 4.11 | 1.78-9.51 | **0.001** |
| **Cough for over 3 weeks** | | | | | | |
| **No** | Ref. | | | Ref. | | |
| **Yes** | 1.29 | 0.44- 3.79 | 0.641 | 2.04 | 0.97-4.29 | 0.061 |
| **Night sweats[b, d]** | | | | | | |
| **No** | Ref. | | | | | |
| **Yes** | 8.25 | 1.15-59.47 | **0.036** | | | |
| **Constitutional syndrome (asthenia anorexia weight loss)** | | | | | | |
| **No** | Ref. | | | Ref. | | |
| **Yes** | 2.07 | 0.66-6.45 | 0.211 | 2.94 | 0.44-19.80 | 0.268 |

[a] Client participants tested for HIV in both observation phases. 14 individuals with an undetermined HIV test result were excluded. For the multivariable analysis, 1471 men and 2422 were included, respectively.

[b] Sociodemographic characteristics, risk factors or signs and symptoms not included in the national targeted PITC algorithm of Mozambique.

[c] Occasionally: less than half of the times; Frequently: more than half of the times

[d] The aOR among women could not be estimated because there was not any woman included in the multivariable analysis who presented with night sweats.

Abbreviations: aOR: adjusted odds ratio, CI: confidence interval, PITC: provider-initiated testing and counselling, Ref.: reference category
